# Determinants of COVID-19 Incidence and Mortality in the US: Spatial Analysis

**DOI:** 10.1101/2020.12.02.20242685

**Authors:** Niranjan J. Kathe, Rajvi J. Wani

## Abstract

**OBJECTIVES:** The US continues to account for the highest proportion of the global Coronavirus Disease-2019 (COVID-19) cases and deaths. Currently, it is important to contextualize the spread and success of mitigation efforts. The objective of this study was to assess the ecological determinants (policy, health behaviors, socio-economic, physical environment, and clinical care) of COVID-19 incidence and mortality in the US.

**METHODS:** Data from the New York Times’ COVID-19 repository (01/21/2020-10/27/2020), 2020 County Health Rankings, 2016 County Presidential Election Returns, and 2018-2019 Area Health Resource File were used. County-level logged incidence and mortality rate/million were modeled using the Spatial Autoregressive Combined model and spatial lag model.

**RESULTS:** Counties with higher proportions of racial minorities (African American β= 0.007, Native Americans β= 0.008, Hispanics β= 0.015), non-English speakers (β= 0.010), population density ([logged] β= 0.028), and air pollution (β= 0.062) were significantly associated with high COVID-19 incidence rates. Subsequently, counties with higher Republican voters (β= 0.017), excessive drinkers (β= 0.107), children in single-parent households (β= 0.018), uninsured adults (β= 0.038), racial minorities (African American β= 0.032, Native Americans β= 0.034, Hispanics β= 0.037), females (β= 0.101), and population density ([logged] β= 0.270), air pollution (β= 0.130), and non-Whites/Whites’ residential segregation (β= 0.014) were significantly associated with high COVID-19 mortality rates. Additionally, longer state-level restrictions were associated with lower COVID-19 incidence and mortality rates.

**CONCLUSIONS:** The spatial models identified longer state-level restrictions, population density, air pollution, uninsured rate, and race/ethnicity as important determinants of the geographic disparities in COVID-19 incidence and mortality.

## INTRODUCTION

The Coronavirus Disease 2019 (COVID-19) pandemic continues to spread in the US and around the world. The US, as of January 11, 2021 recorded 22.6million COVID-19 cases and 375,141deaths. ^1^ The US cases and deaths continue to account for the largest share of global cases (25%) and global deaths(20%).^1^ The COVID-19 pandemic containment in the US was challenging actors due to virus contagion characteristics, its pathophysiology, and socio-political factors. In response to the pandemic, by the first week of April, all but two states and local authorities imposed social distancing measures and restrictions. As of the writing of this study, many states who had eased phase 1 restrictions went on to implement phase 2 restriction policies to curb the surge in COVID-19 infections during the holiday season. ^2–5^

Restriction measures such as the closure of business establishments, stay-at-home, and social distancing mandates severely impacted the economy. In response to the pandemic, the lawmakers passed the Coronavirus Aid, Relief, and Economic Security Act (CARES) Act, ^6^ with additional relief expected shortly. As of January 11, 2021, Moderna and Pfizer vaccines have received emergency use authorization by the FDA and are rolled out in phases. Other biopharmaceutical companies are conducting clinical trials for fifty-four COVID-19 vaccine candidates.^7–9^ The current vaccine rollout will continue to gain momentum while other COVID-19 vaccines may receive market approval in the near future. Despite this progress, experts recommend that the public follow COVID-19 safety measures due to the slow initial rollout and uncertainties surrounding COVID-19 vaccines (virus mutations, duration of immunity, real world effectiveness, vaccine uptake, etc.).^10^ In summary, fiscal, legislative, and scientific efforts are all underway to address the needs of the population in the current phase of the COVID-19 pandemic. However, the assessment of the impact of ecological contextual factors such as health behaviors, clinical care, socio-economic, and physical environment-related characteristics on the course of COVID-19 pandemic is also necessary. The contextual understanding from such a study is required to gauge which strategies work, to what extent, and for which groups. Having a thorough knowledge of these ecological contextual factors is critical to address the public health and economic challenges and prioritize resources.

Studies so far have generated predictive models for growth in COVID-19 incidence and mortality and estimated the impacts of some community-level factors on COVID-19 incidence and mortality. Millett *et al*. and Khanijahania *et al*. focused on assessing the ecological determinants of susceptibility to COVID-19 outcomes among predominantly African American counties, while Fielding-Miller *et al*. and Peters *et al*. performed a similar assessment along the rural-urban continuum.^11–14^ Few studies had assessed the spatial determinants of COVID-19 transmission.^15–17^ Recently, Sun *et al*. assessed the relationship between various county-level determinants and COVID-19 incidence while Andersen *et al*. also identified high prevalence clusters.^15,16^ These studies had evaluated the incidence and prevalence during the initial phase of the pandemic and limited the analysis to a few county-level factors. The goal of the current study was to assess the impact of county-level ecological factors, using spatial econometric analysis, on the cumulative COVID-19 incidence and mortality rate over an extended phase of the pandemic.

## METHODS

### Data source and study design

The current study used county-level COVID-19 confirmed cases and deaths data from the New York Times repository extracted as of October 27, 2020, and included data up until that date.^18^ The study used county-level characteristics from 2020 County Health Rankings data, 2018-2019 Area Health Resource File data, 2016 County Presidential Election Returns data, and state-level lockdown duration data was generated based on the lockdown start and end date obtained from Ballotpedia as of October 27, 2020 ^19–21^. Counties from Alaska and Hawaii and counties with less than 25 cumulative cases as of October 27, 2020, were excluded from the analysis. The US counties ESRI Shapefile were obtained from the US Census Bureau.^22^ The study employed a cross-sectional ecological study design to assess the association between county-level characteristics on the cumulative COVID-19 incidence cases and deaths.

### Outcomes

The county-level cumulative COVID-19 cases and COVID-19 deaths as of October 27, 2020 (per million population) were the operational definition for COVID-19 incidence and mortality rate, respectively. The resulting incidence and mortality rates were log-transformed due to the skewed nature of the county-level cumulative incidence and mortality rates.

### Covariates

The covariates selected to predict the county-level incidence and mortality were based on the County Health Ranking framework. The framework categorizes health factors into four sub-categories, namely health behaviors, clinical care, socioeconomic factors, physical environment (**Figure 1**). Each of the sub-categories is further divided into individual components. Although the County Health Ranking model assigns weights to each of the components, they were not utilized in the current analysis as no composite rank score was calculated. The individual components were used as covariates in the regression. The final list of county-level covariates included in the model is described in **Supplementary Table 1**. In addition to the above county-level factors, length of state-level lockdown duration and month (January/February/March) of the first reported case at the state-level were included as covariates. Finally, the county-level composition of the Republican voters was also added to determine the extent to which the county’s community health was associated with voting preference for the 2016 presidential elections.

**Figure 1:**
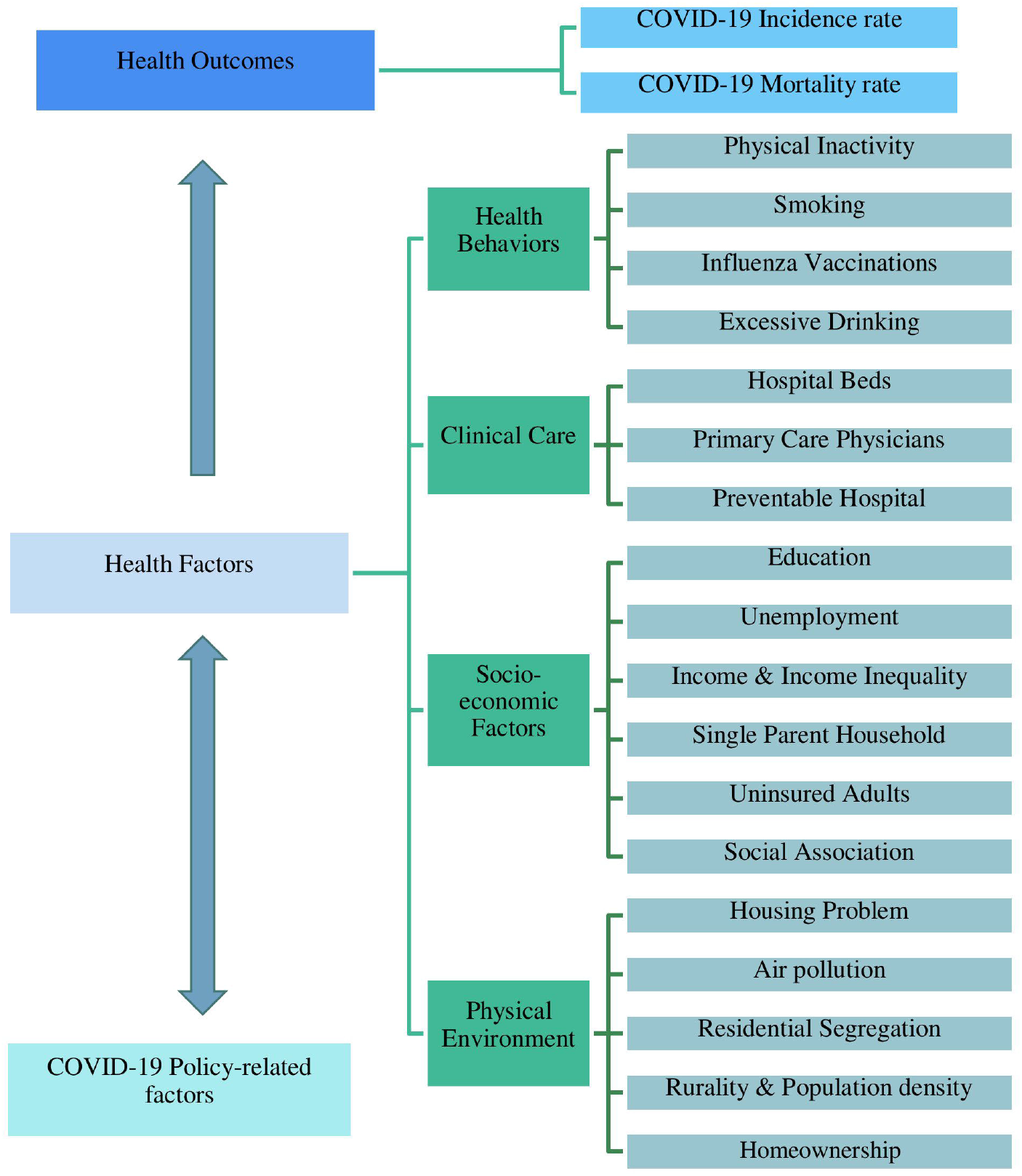
Theoretical framework based on the county health rankings model to establish a relationship between COVID-19 incidence and mortality, and county-level ecological factors.

### Statistical Analyses

Descriptive univariate statistics of the weighted county-level characteristics were generated. The presence of spatial correlation was confirmed by performing Moran’s I test for correlation of ordinary least square regression residuals. Due to the spatial correlation in the data, the current study employed a spatial regression analysis approach. Two island counties were excluded from the spatial regression analysis. Prior research on spatial analysis of COVID-19 used the spatial autoregressive combined model (SAC) model to evaluate the association between period prevalence of COVID-19 and county-level characteristics ^15^. The SAC model was also adopted for modeling cumulative incidence and deaths if both the spatial lag parameter (rho) and spatial error parameter (lambda) were statistically significant. However, model simplification was attempted when one of the parameters was not significant. A first-order queen spatial weight matrix was employed for all spatial models. The queen matrix defines neighbor relationships if the counties either share a border or a vertex. All analysis was performed in SAS Studio University Edition (Cary, NC), QGIS v 3.16.0 (Berne, Switzerland), and RStudio (R) v 4.0.3 (Boston, Massachusetts).

## RESULTS

The final analysis included data from 3,101 counties from the mainland US. Between January 20 to October 27, 2020, the population-weighted cumulative incidence and mortality rates for the mainland US were 26,576 and 632 per million, respectively. **Table 1** demonstrates descriptive statistics of the incidence and mortality rates and county-level determinants, namely, health behaviors, clinical care, socio-economic, and physical environment factors. Fifteen states had a length of the lockdown of 59 days or more, and 39 states reported their first case in the month of March. Overall, 2,499 counties leaned Republican (50% of the votes in the county were for the Republican presidential candidate in 2016). The weighted proportion of adult smokers, adults with obesity, adults with physical inactivity, Medicare enrollees that were administered influenza vaccines and adults with some college education were 15%, 29%, 23%, 19%, and 65%, respectively. Premature (<75 years) age-adjusted mortality was 344 per 100,000. Among socio-economic factors, unemployment was 4%, 12% of adults were uninsured, the mean income inequality ratio was 5. About 12% were African Americans, 18% Hispanics, 1% Native Americans, and 4% population not proficient in English. The mean percentage of the population older than 65 years and less than 18 years were 16% and 23%, respectively. On average, 19% of counties were rural, the homeownership rate was 65%, 33% of the children lived in single-parent households, and 18% of households had severe housing problems. At the county-level, the rate of primary care physicians (logged) and preventable hospitalization (logged) was 4 and 8, respectively. Importantly, no significant multicollinearity was observed in the analysis, as the VIF for the selected variables was less than 7.

**Table 1:** Descriptive statistics of factors used in this study, as of October 27, 2020 (n = 3,101) using AHRF, NYT, CPER and CHR datasets.

| Ecological Factors | Mean | SD |
| --- | --- | --- |
| Incidence rate in per 1 Million (logged) | 10.07 | 0.53 |
| Incidence rate in per 1 Million | 26,575.75 | 12,216.59 |
| Mortality rate in per 1 Million (logged) | 6.18 | 0.80 |
| Mortality rate in per 1 Million | 631.59 | 477.64 |
| **COVID-19 Policy- related factors** |  |  |
| Length of order (quintiles) [n, %]* | *n* | *%* |
| 0 days (No order) | 438.00 | 14.12 |
| 1 - 27 days | 469.00 | 15.12 |
| 28 - 35 days | 717.00 | 23.12 |
| 36 - 58 days | 714.00 | 23.02 |
| 59 or more days | 736.00 | 24.60 |
| Calendar month of the first reported case in the state [n, %]* | *n* | *%* |
| January | 214.00 | 6.90 |
| February | 497.00 | 16.03 |
| March | 2,390.00 | 77.07 |
| **Health behaviors** |  |  |
| % Adult Smoking | 15.36 | 3.46 |
| % Physical Inactivity | 23.25 | 5.37 |
| % Excessive Drinking | 18.76 | 2.79 |
| % Influenza Vaccinations^ | 46.27 | 6.91 |
| **Clinical Care** |  |  |
| Primary Care Physicians Rate (log) | 4.23 | 0.51 |
| Preventable Hospitalization Rate (log) | 8.38 | 0.29 |
| Hospital Beds (log) | 6.45 | 3.29 |
| **Socio-economic factors** |  |  |
| % Republicans | 46.63 | 16.77 |
| Counties leaning Republican (>50% votes) [n, %] | 2,500.00 | 80.62 |
| % Adult Obesity | 29.23 | 5.45 |
| Premature Age Adjusted Mortality<sup>#</sup> | 343.74 | 92.96 |
| Income Inequality Ratio | 4.67 | 0.68 |
| % Adults with Some College Education | 65.28 | 9.79 |
| % Unemployment | 3.95 | 1.13 |
| % Children in Single-parent Households | 32.99 | 8.58 |
| Social Association Rate | 9.09 | 3.57 |
| % Uninsured Adults | 12.42 | 6.00 |
| Median Income per \$1,000 (log) | 4.14 | 0.26 |
| % 65 years old and over | 16.05 | 4.08 |
| % less than 18 years of age | 22.49 | 2.91 |
| % African American | 12.30 | 12.56 |
| % Native Americans | 1.23 | 3.22 |
| % Hispanic | 18.06 | 17.19 |
| % Female | 50.72 | 1.22 |
| % Not proficient in English | 4.19 | 4.20 |
| **Physical environment** |  |  |
| Air pollution-particulate matter (2.5) | 10.09 | 2.13 |
| % Severe Housing Problem | 17.72 | 5.41 |
| Residential Segregation- African Americans/White | 50.67 | 12.82 |
| Residential Segregation- non-White/White | 36.73 | 11.25 |
| % Rural | 19.18 | 24.18 |
| % Homeownership | 64.57 | 9.54 |
| Population density per 100 sq. miles (log) | 6.12 | 1.55 |

Abbreviations: AHRF: 2018-2019 Area Health Resource Files; 2020 NYT: New York Times; 2020 CHR: County Health Rankings; and CPER: 2016 County Presidential Election Returns; SD: Standard Deviation.
\* Unweighted as this represents the number of counties; ^: Among Medicare enrollees; #: Number of deaths among residents under age 75 per 100,000 population (age- adjusted).

**Figure 2** presents the spatial distribution(septiles) of logged COVID-19 incidence rates. In the West, Oregon High desert and Twin Falls areas, New Mexico’s Navajo and Apache areas, and Washington’s Spokane areas had clusters of high incidence rates. In the Midwestern region, Tribal lands and the Great Plains regions, including North Dakota, South Dakota, Nebraska and Iowa (congruence of these borders near Sioux Falls area), Minnesota, Wisconsin (specifically Superior Upland area), and Kansas regions had clusters with high incidence rate clusters. In the South, the Delta region along the Mississippi river including states of Mississippi, Louisiana, Tennessee, central Alabama, and Arkansas had clusters of high incidence rates. The Corpus Christie area and area along the US-Mexico border had clusters of high incidence rates clusters. Additional high incidence rate clusters were found in the Tallahassee, Jacksonville, Savannah area, and Miami-Dade County area. Conversely, low incidence rates were in the Pacific-Northwest areas. The spatial distribution of the logged COVID-19 mortality rates (by septiles) presented in **Figure 3** demonstrated similar patterns. However, in the Northeastern region, high mortality rate clusters were found between the greater Philadelphia area and Boston Massachusetts. Contrarily, the mortality rate clusters in Wisconsin’s Superior Upland area were smaller compared to the incidence rate clusters.

**Figure 2:**
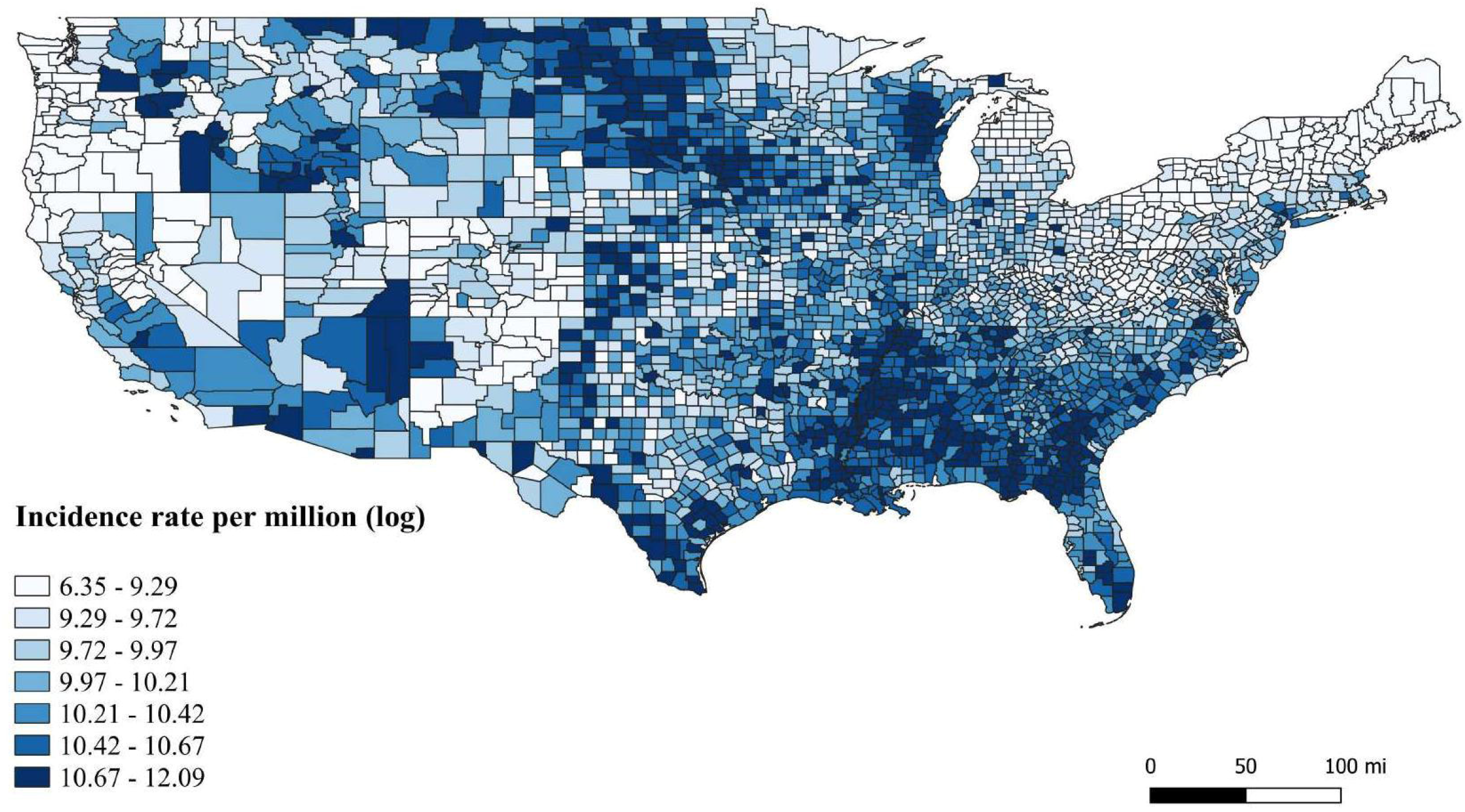
Spatial distribution of the logged cumulative COVID-19 incidence rates by septiles, as of October 27, 2020.

**Figure 3:**
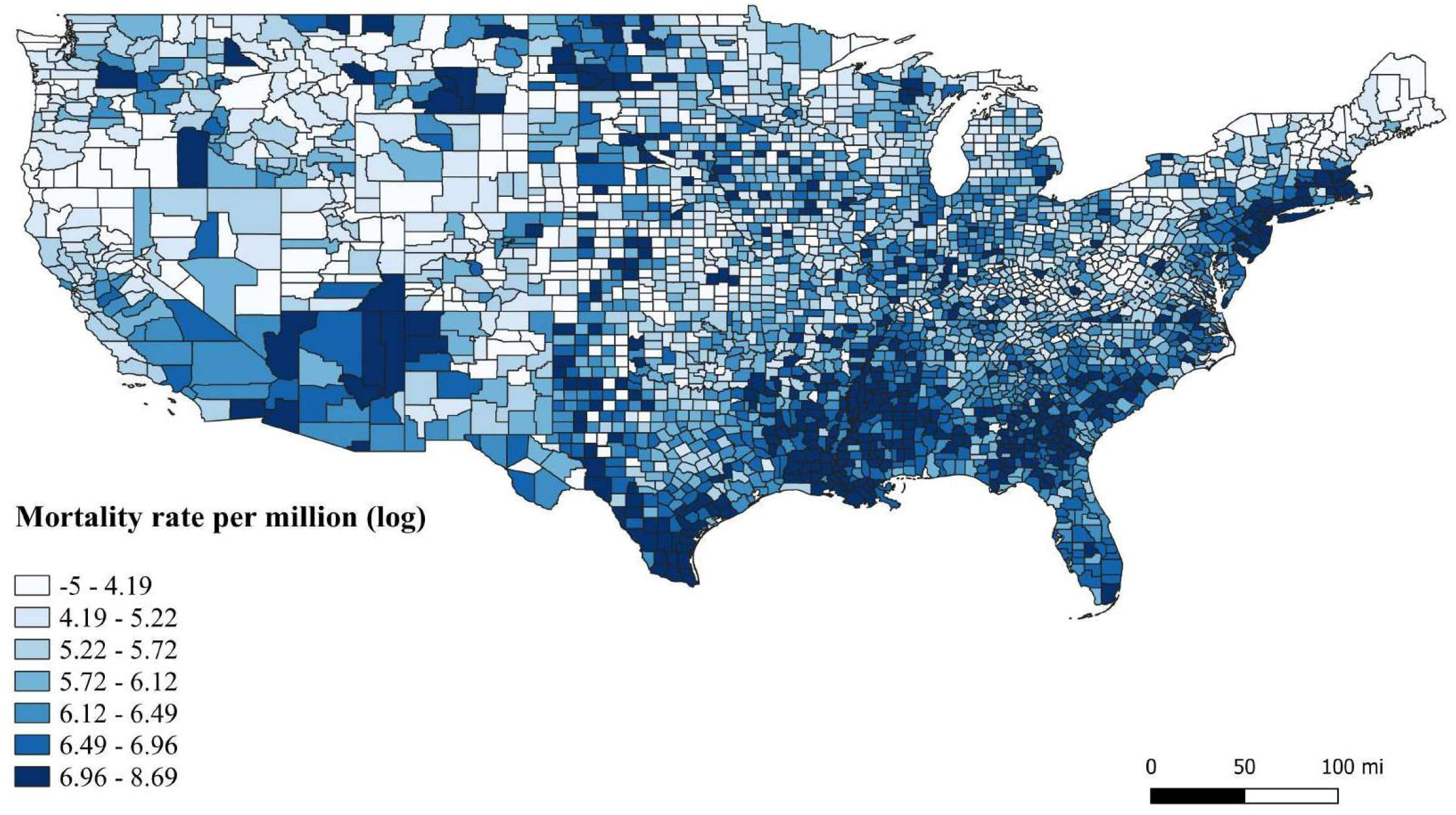
Spatial distribution of the logged cumulative COVID-19 mortality by septiles, as of October 27, 2020.

**Table 2** presents the results from spatial regression models that assessed the impact of ecological determinants on COVID-19 incidence and mortality rates (logged). For the incidence rate SAC regression, the rho and lambda model parameters both were significant. However, only the rho parameter was significant for the mortality rate regression model. Therefore, the SAC model was used for incidence rate analysis, while the spatial lag model was used for mortality rate analysis. The analysis further tested the spatial correlation for the residuals of both models using Moran’s I statistic. The Moran’s I for the model residuals was not significant for the incidence rate (SAC model) and mortality rate (spatial lag model) regression, which indicated no significant spatial autocorrelation of residuals. The length of lockdown order, a policy-related factor, was significantly associated with the incidence rate. Compared to the counties that had no orders implemented, counties with an order length of 1-27 days, 36-58 days, and 59 or more days had a 0.290, 0.228, and 0.335 unit decrease in logged COVID-19 incidence rates, respectively. Among clinical care-related factors, a unit increase in logged primary care physicians’ rate, logged preventable hospitalization rates and logged hospital beds were all associated with 0.011, 0.045, and 0.004 unit increase in logged COVID-19 incidence rates, respectively. A percentage point increase in influenza vaccinations and Republican political leaning was associated with a 0.003 and 0.002-unit increase in logged COVID-19 incidence rates, respectively. Among other socio-economic factors, a percentage point increase in adults with some college education, children in single-parent households, and social association rate and logged median income resulted in 0.004, 0.003, 0.003, and 0.158 unit decreases in logged incidence rates, respectively. Specifically, among the racial/ethnic factors, a percentage point increase in African Americans, Native Americans, Hispanics, and those not proficient in English was associated with 0.007, 0.008, 0.015, and 0.010-unit increase in logged incidence rates, respectively. Conversely, a percentage point increase in 65 years or older and those less than 18 years of age was associated with a 0.020 and 0.011 unit decrease in logged incidence rates. A unit increase in African American and white residential segregation and a percentage point increase in rurality were associated with a 0.002 and 0.001 unit decrease in logged incidence rates. On the other hand, a unit increase in white and non-white residential segregation and particulate matter air pollution were associated with an 0.003 and 0.062-unit increase in incidence rates, respectively.

**Table 2:**
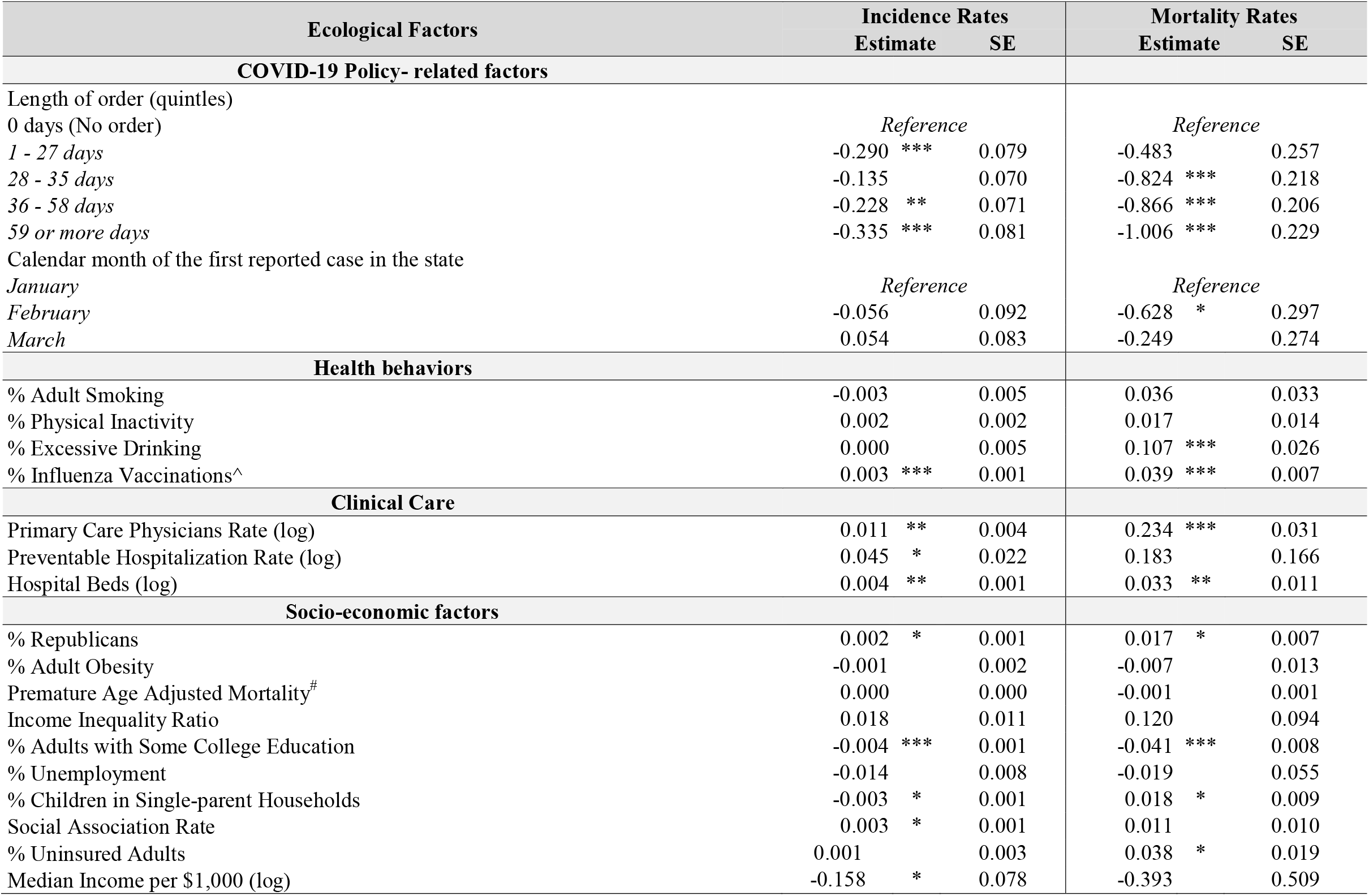
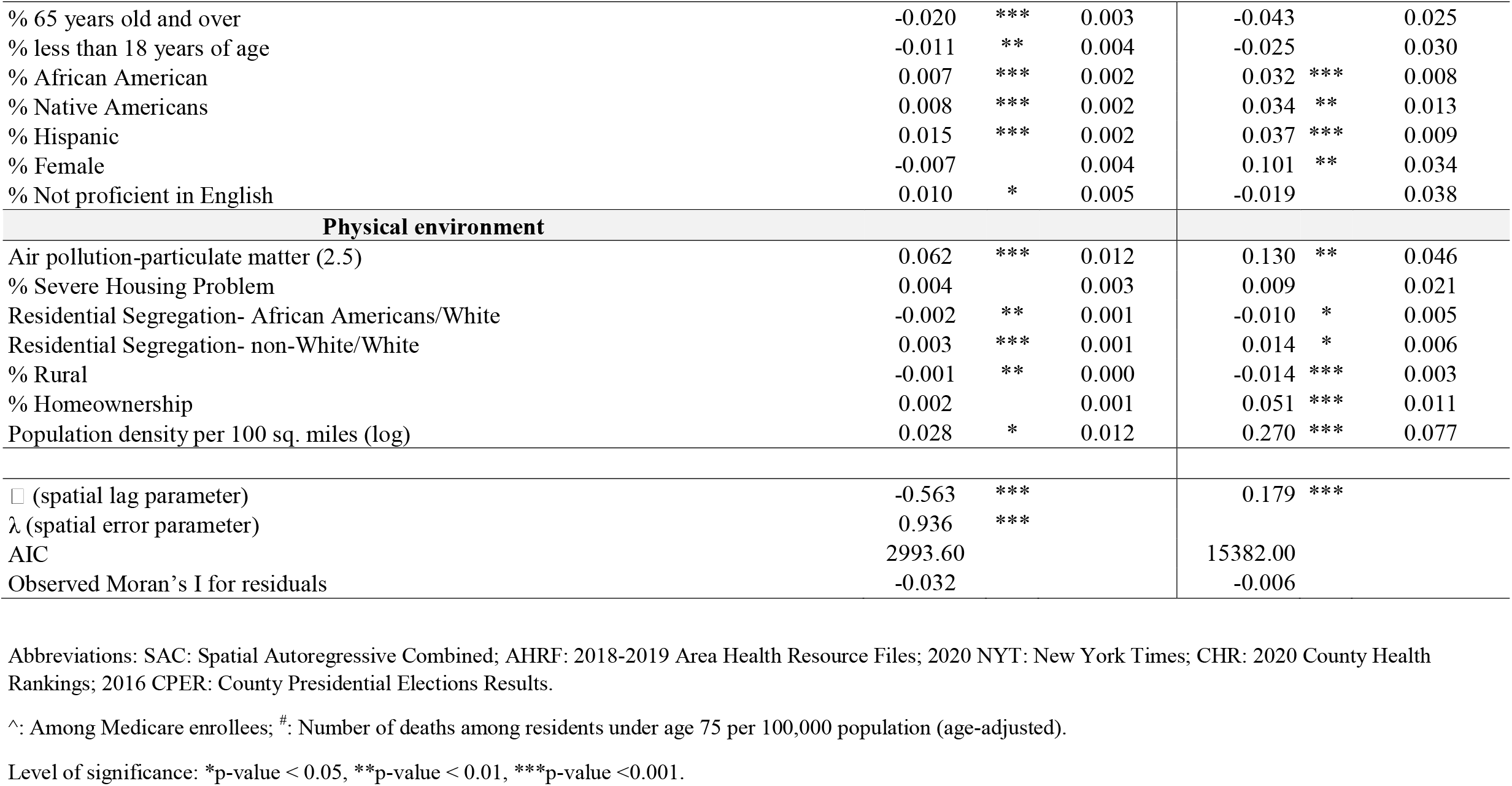
SAC model for the incidence rates (logged) and Spatial Lag Model for mortality rates (logged) as of October 27, 2020 using NYT, AHRF, CPER and CHR datasets.

Subsequently, when compared to the counties that had no orders implemented, counties with order length of 28-35 days, 36-58 days, and 59 or more days had a 0.824, 0.866, and 1.006 unit decrease in logged COVID-19-related mortality rates, respectively. Compared to counties from states that first reported COVID-19 cases in January, those with the first reported case in February were associated with a 0.628-unit decrease in logged mortality rates. Among health behaviors, a 1% increase in excessive drinking, influenza vaccinations resulted in a 0.107 and 0.039 increase in logged mortality rates. Logged primary care physicians’ rate and hospital beds resulted in 0.234 and 0.033 units increase in logged mortality rates. Among socio-economic factors, a 1% increase in Republicans, females, children in single-parent households, and uninsured adults resulted in a 0.017, 0.101, 0.018, and 0.038 units increase in logged mortality rates, respectively. However, a 1% increase in adults with some college education was negatively associated i.e. 0.041 decreases in logged mortality rates. Among racial/ethnic composition, a 1% increase in African Americans, Native Americans, and Hispanics were associated with 0.032, 0.034, and 0.037 unit increase in logged mortality rates. The logged population density was also positively associated with incidence and mortality rates by 0.028 and 0.270-units, respectively. Additionally, a 1% increase in air pollution, and residential segregation-Non-White/Whites resulted in a 0.130 and 0.014 increase in logged mortality rates. Whereas African Americans/White residential segregation, and rurality were negatively associated with mortality rates by 0.010 and 0.014 units.

## DISCUSSION

To the best of our knowledge, this is the first spatial analysis study that captured and assessed the cumulative incidence and deaths during the majority of the year 2020 (January 21 to October 28, 2020) of the COVID-19 pandemic in the US. The study found that counties with higher proportions of Republican voters, social association rates, racial minorities (African Americans, Native Americans, and Hispanics), those not proficient in English and counties with higher residential segregation between non-Whites & Whites, population density, pollution-particulate matter were associated with higher incidence rates. However, counties with longer length of stay-at-home orders, a higher proportion of adults with some college education, high-income, elderly, children, rurality, and higher segregation between African Americans & Whites had lower incidence rates. Correspondingly, counties with higher Republican voters, excessive drinkers, children in single-parent households, uninsured adults, racial minorities (African Americans, Native Americans, and Hispanics), females, and higher population density, pollution-particulate matter, and residential segregation between non-Whites & Whites had higher COVID-19 mortality rate.

Few studies have assessed the impact of different ecological factors on COVID-19 incidence and mortality rates in the US and worldwide. Similar to the current study, Peters *et al*. reported higher age, population density, uninsured rate to be associated with increased susceptibility to COVID-19 outcomes ^13^. Khazanchi *et al*. and Nayak *et al*. also reported a similar association between higher county-level susceptibility score and higher incidence of COVID-19 incidence and deaths ^12,23^. Similar to our study, Liang *et al*. also assessed the impact of air pollution level and found increased air pollution to be associated with an increased mortality rate ^24^. While Fielding-Miller *et al*. reported higher COVID-19 mortality for counties with higher non-English speaking populations, our study showed higher incidence rates among this group ^14^. Also, in agreement with the current study, other studies have reported high incidence and mortality rates in counties with greater proportions of racial minorities (Native Americans, Hispanics, and African Americans) ^12,15,25^. Moreover, with an increase in population density, incidence and mortality rates increased, which has been reported by Sun *et al*. Unlike this study, Mollalo *et al*. spatial analysis found that increase in household income was associated with an increased incidence rate ^26^. However, that same study reported that an increase in providers (for example nurse practitioners) increased incidence rates, which was similar to our findings. Allcott *et al*. and Goolsbee *et al*. also have pointed out ‘partisanship’ as a risk factor for non-adherence to preventive guidelines and mask use. Our study also observed that higher Republican-leaning counties were associated with higher incidence and mortality ^27,28^. Interestingly, similar to Sun *et al*., the current study also reported a lower incidence among counties with higher proportions of people over 65 years of age ^15^.

The study has important limitations. First, log transforming the outcomes i.e. cumulative incidence and mortality rates as a linear dependent variable may mask the variations across counties. To our understanding, there is no software package currently available that runs a spatial lag model with a dependent variable with Poisson or Binomial distribution and thus, this study transformed the outcomes in their logarithmic form. Given the cross-sectional nature of the study, no causal inferences can be made. There are considerable differences in the testing rates across regions and counties and can influence the observed incidence rate. The list of variables is by no means comprehensive and does not include several other factors such as mobility, local restriction policies (county or city-level), compliance with local and federal prevention guidelines. The current analysis is ecological in nature and no direct inferences can be drawn at the individual level.

## CONCLUSION

The findings of this study are more insightful than the mere coronavirus count meters and data visualizations that depict the spread of the COVID-19 pandemic. The current spatial models incorporated a comprehensive list of factors to ensure that the results, when parsed, offer a multi-faceted explanatory power. For illustration, these models helped identify factors including policy-related factors (i.e. length of order), health behaviors (example: excessive drinking), clinical care (example: preventable hospitalization rate), socio-economic factors (example: race/ethnicity, median income, uninsurance rate, education), and physical environment (example: population density, air pollution-particulate matter, rurality, residential segregation among races, homeownership) as some of the important determinants of the geographic disparities in COVID-19 incidence and mortality. As per prior research, this study reaffirms that policy restrictions have helped to limit the COVID-19 incidence and mortality. This study highlights the plausible effect of one’s residential location, vicinity, and local policymakers; and the connectivity to the neighboring counties on the incidence and mortality of COVID-19. As the country is facing the next wave of pandemic, the study findings have important policy implications and guidance on identifying areas at greater risk of infection and mortality. As the pandemic gains momentum in the rural areas, especially in the Midwest and South, contextualized policies at the local level that align with state and federal policies will be necessary to contain the next wave.

## Data Availability

County-level COVID-19 confirmed cases and deaths data from the New York Times is publicly available.

https://github.com/nytimes/covid-19-data

