## Supplementary Table 1 for "Determinants of COVID-19 Incidence and Mortality in the US: Spatial Analysis"

**Supplementary Table 1: Definition of the ecological factors used in the analysis, their source definitions and the modifications.**

| **Factors** | **Source Definition** | **Modification** |
| --- | --- | --- |
| Log of Cumulative COVID-19 incidence rates | Total number of reported COVID-19 cases between 20st Jan 2020 and 28th October 2020 per 1 million population | Log (Source variable + (10)^-6^); categorized into septlies for the maps |
| Log Cumulative COVID-19 mortality rates | Total number of reported COVID-19 deaths between 20st Jan 2020 and 28th October 2020 per 1 million population | Log (Source variable + (10)^-6^); categorized into septlies for the maps |
| Length of order | Number of days for which the stay-at-home, shelter-in-place, safer at home, or other such safety orders implement. This variable was divided into five categories 0 days, 1-27 days, 28-35 days, 36-58 days and 59 or more days |  |
| Calendar month of the first reported case in the state | Month of the year when the first COVID-19 case was reported in the state. This variable fell into three categories-January, February and March. |  |
| Percentage Republicans | Percentage of people who votes for the Republican party in 2016 |  |
| Percent Adult smokers | Percentage of adults who are current smokers |  |
| Percent Adult Obese | Percentage of the adult population (age 20 and older) that reports a body mass index (BMI) greater than or equal to 30 kg/m^2^ |  |
| Percent excessive drinking | Percentage of adults reporting binge or heavy drinking |  |
| Percent flu vaccine | Percentage of fee-for-service (FFS) Medicare enrollees that had an annual flu vaccination |  |
| Age adjusted mortality | Number of deaths among residents under age 75 per 100,000 population (age-adjusted) |  |
| Percent physical inactivity | Percentage of adults age 20 and over reporting no leisure-time physical activity |  |
| Income inequality ratio | Ratio of household income at the 80th percentile to income at the 20th percentile |  |
| Log PCP rate | Log of Ratio of population to primary care physicians | Log (Source variable + (10)^-6^) |
| Log preventable hospitalization rate | Rate of hospital stays for ambulatory-care sensitive conditions per 100,000 Medicare enrollees | Log (Source variable + (10)^-6^) |
| Percent Educated at some college | Percentage of adults ages 25-44 with some post-secondary education |  |
| Percent unemployment | Number of people ages 16 years and above unemployed and looking for work |  |
| Percent single parent | Percentage of children that live in single-parent households |  |
| Social Association rate | Number of membership associations per 10,000 population |  |
| Pollution Level: PM 2.5 | Average daily density of fine particulate matter in micrograms per cubic meter (PM2.5) |  |
| Percent problem in housing | Percentage of households with at least 1 of 4 housing problems: overcrowding, high housing costs, lack of kitchen facilities, or lack of plumbing facilities |  |
| Percent uninsured adult | Percentage of adults under age 65 without health insurance. |  |
| Median income in thousands | The income where half of households in a county earn more and half of households earn less | Source variable /1000 |
| Percent over 65 | Percentage of population ages 65 and older |  |
| Percent less than 18 | Percentage of population below 18 years of age |  |
| Percent Black | Percentage of population that is non-Hispanic Black or African American |  |
| Percent Native American | Percentage of population that is American Indian or Alaska Native |  |
| Percent Hispanic | Percentage of population that is Hispanic |  |
| Percent female | Percentage of population that is female |  |
| Segregation index: White/Black | Index of dissimilarity where higher values indicate greater residential segregation between Black and White county residents |  |
| Segregation index: White/non-White | Index of dissimilarity where higher values indicate greater residential segregation between non-White and White county residents |  |
| Percent Rural | Percentage of population living in a rural area |  |
| Log population density | Persons per square mile area | Log (source variable) |
| Percent not proficient in English | Percentage of population that is not proficient in English. |  |
| Percent homeownership | Percentage of occupied housing units that are owned. |  |
| Log of hospital bed rates^#^ | Number of beds regularly maintained (set up and staffed for use) for inpatients as of the close of the reporting period | Log (Source variable + (10)^-6^) |
| Log of population | Resident population | Log (source variable) |

**Note**:

Variables obtained from AHRF: 2018-2019 Area Health Resource Files; 2020 NYT: New York Times; 2020 CHR: County Health Rankings; and CPER: 2016 County Presidential Election Returns

^#^ Variables obtained from Area health resource File
